# Search for asymptomatic carriers of SARS-CoV-2 in healthcare workers during the pandemic: a Spanish experience

**DOI:** 10.1101/2020.05.18.20103283

**Authors:** Julián Olalla, Ana M Correa, M Dolores Martín-Escalante, M Luisa Hortas, María Jesús Martín-Sendarrubias, Víctor Fuentes, Gabriel Sena, Javier García-Alegría, on behalf the ROBLE group

**Affiliations:** Internal Medicine Unit, Hospital Costa del Sol, Marbella, SPAIN; Microbiology Unit, Hospital Costa del Sol, Marbella, SPAIN; Laboratory Area, Hospital Costa del Sol, Marbella, SPAIN; Preventive Medicine, Hospital Costa del Sol, Marbella, SPAIN; Occupational Health Department, Hospital Costa del Sol, Marbella, SPAIN

## Abstract

**Objective:** determine the percentage of healthcare workers (HCW) carrying SARS-CoV-2 in high exposure areas of the hospital.

**Design:** cross-sectional study during April 15–24^th^ in Hospital Costa del Sol (Marbella, Spain), excluding HCW with previous COVID19.

**Setting:** hospital based, focused on patient care areas COVID19.

**Participants:** 498 subjects, 80% women. Participation was offered to all the HCW of Emergencies, Intensive Care and Anesthesia, Internal Medicine and Pneumology. Other units not directly involved in the care of these patients were offered to participate.

**Intervention:** naso and oropharyngeal PCR determination was performed together with IgG and IgM antibody determination by immunochromatography. On the day of sampling, a health questionnaire was answered, reporting symptoms on the same day and in the previous fourteen days.

**Main outcome measures:** percentage of HCW with positive PCR for SARS-CoV-2, percentage of HCW with positive IgG for SARS-CoV-2.

**Results:** Two individuals were detected with PCR for SARS-CoV-2 positive (0.4%). Both were asymptomatic on the day of sampling, but one of them had had a CoVID-19 compatible picture in the previous two weeks and had positive IgG and IgM; therefore, only one subject was truly asymptomatic carrier (0.2%). 9 workers with positive IgG (1.8%) were detected.

**Conclusions:** the prevalence of asymptomatic carriers among health workers of the services directly involved in the care of patients with CoVID-19 was very low in our center. This type of strategy can be one more tool in controlling the pandemic.

---

At the end of January 2020, the first case of SARS-CoV-2 infection was declared in Spain(1). In China’s initial outbreak, health workers had accounted for 3.8% of total reported cases(2), but in Spain, at the end of April 2020 this group accounted for 20% of all cases(3). From the beginning of the pandemic, the identification of asymptomatic subjects carrying SARS-CoV-2 has been advocated as one of the fundamental measures to reduce its spread(4). The importance of these subjects is that they can act as vehicles for the virus, transmitting it to individuals who may be infected. Models have been carried out that have attempted to estimate the proportion of asymptomatic individuals within China, estimating that 86% of all infections were not documented in the first moment of the pandemic, before mobility restrictions, being these responsible for the 79% of documented infections(5).

Therefore, it is important to determine the degree of asymptomatic carriers among the population of health workers, with a view to reducing transmission to other workers and to patients treated for reasons other than CoVID-19. This identification, together with the appropriate measures, could result in less spread of the virus from the healthcare center and, therefore, in fewer healthcare providers and patients affected by COVID-19.

In this article, we present the results of an active search study of asymptomatic and seroprevalence carriers of SARS-CoV-2 among high-risk healthcare workers in a hospital in southern Spain.

## Methods

### Study design

Cross-sectional study carried out between April 15 and 24, 2020 among health workers at the Costa del Sol Hospital in Marbella, in the Autonomous Community of Andalusia (Spain). The study was proposed to the health workers most exposed to direct patient care CoVID-19: Emergencies, Intensive Care (constituted in the crisis situation by personnel from the Intensive Care Unit and Anesthesia and Reanimation), Internal Medicine and Pneumology (plants conventional hospitalization where CoVID patients were admitted). Participation in the study was also offered to the Nephrology, Digestive and Cardiology Units, considering them as non-CoVID plants. When the personnel of a certain area began to attend hospitalized CoVID patients, they were considered for the purposes of this study as members of the Internal Medicine and Pneumology area.

Voluntary participation was offered to all the members of the referred Units, classifying their professional category in: doctors, nurses, nursing assistants, security guards, administrative and cleaning staff. Workers who had been previously diagnosed with CoVID-19 were excluded from the study.

All the members signed an informed consent and authorization was obtained from the management of the center and the approval of the local Ethics Committee to carry it out.

### Procedures

Naso-oropharyngeal exudates and venous blood samples were taken from each professional. The presence of SARS-CoV-2 was detected in respiratory samples using reverse transcriptase polymerase chain reaction (PCR) protocols from 2 providers (VIASURE SARS-CoV-2 from CerTest Biotec and LightMix Modular SARS-CoV (COVID19), Roche). RT-PCR tests followed the manufacturer’s protocols. IgG and IgM antibodies were detected in serum by solid phase immunochromatography (COVID-19 IgG / IgM Rapid Test Cassette, (Zhejiang Orient Gene Biotech Co., LTD).

On the day of the extraction of samples, a health questionnaire was filled in, informing about the presence of a personal history of interest, symptoms on the same day of the extraction and similar symptoms in the previous 14 days. The collected antecedents were: arterial hypertension, heart failure, ischemic heart disease, chronic lung disease, diabetes mellitus, obesity (body mass index> 29), chronic kidney failure, asplenia, chronic liver disease, neuromuscular diseases, immunodeficiency and pregnancy.

The symptoms for which they were asked both on the day of the extraction and on the previous 14 days were: fever, cough, shortness of breath, runny nose, sore throat, diarrhea, vomiting, headache, myalgia and feeling of general malaise. In addition, they specifically asked about contact with CoVID cases in the worker’s environment, outside of the workplace, in the previous 14 days.

### Objectives

The main objective was to determine the prevalence of asymptomatic carriers of SARS-CoV-2 in healthcare professionals, defined as those individuals with PCR of a positive respiratory sample without presenting symptoms suggestive of CoVID-19 on the day of sampling.

Secondary objectives were the percentage of individuals with isolated positive IgG, isolated positive IgM or positive IgG and IgM, always with negative PCR.

### Statistic analysis

A database was generated in SPSS 15, carrying out a descriptive analysis of the variables. In the case of proportions they are expressed in percentages, while in the case of quantitative variables it is done in means and 95% CI.

## Results

### Characteristics of the participants

A total of 498 healthcare workers were recruited, encompassing all the workers in the Emergency, ICU and CoVID-19 plant areas. The main characteristics are shown in Table 1. The mean age of the participants was 41.5 years (95% CI: 40.8–42.3). Of the total participants, 354 (80%) were women. The area that contributed the most participants was the CoVID-19 plant, with 44% of the total individuals in the sample. The most represented professional category was that of nurses, with 195 (39.2%) participants.

**Table 1.** Main characteristics of the participants

| Variable | N (%) |
| --- | --- |
| Número total de participantes | 498 (100) |
| Personal history |  |
| Obesity | 88 (17.7) |
| Cardiovascular disease | 9 (1.8) |
| Arterial hypertension | 30 (6) |
| Heart failure | 2 (0.4) |
| Ischemic heart disease | 3 (0.6) |
| Chronic obstructive pulmonary disease | 46 (9.2) |
| Mellitus diabetes | 4 (0.8) |
| Chronic renal failure | 1 (0.2) |
| Asplenia | 0 (0) |
| Chronic liver disease | 2 (0.4) |
| Neuromuscular disease | 2 (0.4) |
| Immunodeficiency | 3 (0.6) |
| Pregnancy | 2 (0.4) |
| Professional category |  |
| Doctor | 101 (20.3) |
| Nurse | 195 (39.2) |
| Nursing Assistant | 129 (25.9) |
| Watchman | 35 (7) |
| Administrative | 11 (2.2) |
| Housekeeping | 27 (5.4) |
| Distribution by areas |  |
| Emergencies | 129 (25.9) |
| Intensive care | 63 (12.7) |
| CoVID-19 floor | 219 (44) |
| No CoVID-19 floor | 44 (8.8) |
| Pediatrics | 43 (8.6) |
| Symptoms on the day of sampling |  |
| Fever | 1 (0.2) |
| Cough | 50 (10) |
| Breathing difficulty | 5 (1) |
| Rhinorrhea | 40 (8) |
| Sore throat | 40 (8) |
| Diarrhea | 10 (2) |
| Vomiting | 0 (0) |
| Headache | 55 (11) |
| Myalgia | 10 (2) |
| General discomfort | 14 (2.8) |
| Symptoms in the previous 14 days |  |
| Fever | 9 (1.8) |
| Cough | 92 (18.5) |
| Breathing difficulty | 18 (3.6) |
| Rhinorrhea | 65 (13.1) |
| Sore throat | 101 (20.3) |
| Diarrhea | 45 (9) |
| Vomiting | 2 (0.4) |
| Headache | 116 (23.3) |
| Myalgia | 24 (4.8) |
| General discomfort | 46 (9.2) |

Of the personal history, the most frequent was obesity in 88 (17.7%) workers, followed by chronic obstructive pulmonary disease in 46 (9.2%) and high blood pressure in 30 cases (6%). On the day of sampling, 89% of the participants did not report any symptoms. Among those who reported symptoms, the main one was headache in 55 cases (11%), followed by cough in 50 cases (10%), rhinorrhea and sore throat with 40 cases (8%) for each symptom, only one worker reported fever on the day the samples were taken, which was not observed at the time of collection. In no case were any healthcare workers considered to present a suggestive clinical picture of CoVID-19 on the day of sampling. When asked about the symptoms present in the previous two weeks, headache was again the most frequent with 116 cases (23.3%), followed by sore throat in 101 workers (20.3%) and cough in 92 cases (18.5%).

A total of 19 healthcare workers (3.8%) reported contact with CoVID-19 cases outside the workplace.

### Naso and oropharyngeal PCR results

Two participants with positive SARS-CoV-2 PCR were identified. One of them with positive IgG and IgM serology and the other with negative IgG and IgM. They were two nurses from the Emergency Department without any personal record. They had no symptoms on the day of sampling. The healthcare worker with a positive IgG and IgM pattern reported having had diarrhea, general discomfort, and contact with the CoVID-19 case outside the workplace in the previous fourteen days, so it was probably a paucisymptomatic case with persistent naso CRP positive oropharyngeal. So, only one healthcare worker (0.2%) was a true asymptomatic carrier.

### Serology results

Nine health workers with positive IgG were detected (1.8%), one of them with a positive PCR.

Seven of them (1.4%) had IgG positive with negative IgM and PCR. They included two doctors, a nurse, three nursing assistants, and a warden. By areas, two belonged to the ER, one to Intensive Care, two to the CoVID-19 plant and the other two to the non-CoVID-19 plant. None of them reported a background of interest. On the day of the extraction, only one of them reported rhinorrhea as an isolated symptom. In the previous two weeks, one of them reported having presented cough and runny nose, another diarrhea, another headache and two general malaise. In none of these cases was contact with a CoVID-19 case outside the workplace.

Two other workers had positive IgG and IgM. As already mentioned, one of these workers with positive IgG and IgM serology had a positive SARS-Cov-2 PCR in the naso-oropharynx. The other worker was an intensive care nursing assistant with negative SARS-Cov-2 PCR and had no history of interest or symptoms on the day of the extraction or in the previous fourteen days. Nor did it refer to contact with CoVID-19 cases outside the workplace.

Two more workers (0.4%) presented positive IgM with negative IgG and negative PCR. It was a CoVID-19 floor doctor and a non-CoVID-19 floor nurse, with no relevant history. On the day of extraction, one of them reported headache and in the previous two weeks the other worker reported having presented a cough. After 14 days of sampling, both remained asymptomatic.

## Discussion

In this cut-off study in healthcare workers with high occupational exposure, we have detected a percentage of asymptomatic carriers of SARS-CoV-2 of 0.4%. Actually, one of the two health workers detected as asymptomatic carriers presented a suggestive clinical picture of CoVID-19 in the two previous weeks, which together with the presence of positive IgG and IgM suggests that it was a CoVID-19 case with little clinical expression. Thus, true asymptomatic carriers would decrease to 0.2%, with a single case among 498 workers investigated. The percentage of asymptomatic individuals with positive IgG was 1.8%, excluding the worker who presented positive PCR, this percentage would decrease to 1.6%, without any symptoms suggestive of CoVID-19 being identified in the fourteen days prior to sampling.

The percentage of true asymptomatic carriers is considerably lower than that reported in other healthcare centers in our country, with higher incidence rates than those in southern Spain. In a published study that communicates the results of PCR in respiratory samples for SARS-CoV-2 in a Madrid hospital, it was found that 11.9% of the workers in the total workforce presented positive PCR, although in this study it was performed testing only symptomatic workers(6).

Our hospital treated the first case of CoVID-19 on February 28^th^ and the first ill healthcare worker was diagnosed on March 11. From that day until April 24, a total of 50 health workers have been diagnosed with CoVID-19, constituting 2.27% of the workforce. This difference with respect to the percentage of health workers reported in Madrid and in Spain as a whole, reflects the differences in incidence between Andalusia and the rest of Spain. During the two weeks of the study, the incidence of CoVID-19 cases in Andalusia remained at around 30 cases per 100,000 inhabitants(7).

The active search for asymptomatic carriers constitutes an additional measure that could affect the limitation of nosocomial transmission of SARS-CoV-2. Nosocomial outbreaks have been reported in residences whose door of entry could have been the presence of an infected worker(8) and nosocomial transmission has been presumed in up to 29% of the healthcare workers infected at the start of the pandemic(9). In these outbreaks, the presence of asymptomatic carriers reaches more than half of the individuals with positive PCR, although after a few days the truly asymptomatic ones are very rare. This coincides with our finding that, in a hospital care setting that includes CoVID-19 patients, the proportion of asymptomatic carriers is minimal. This strategy remains one of the weaknesses in the control of the pandemic(10), as long as transmission has been described from asymptomatic cases(11, 12). Other screening strategies based on typical symptoms of CoVID-19 in healthcare workers have shown that about 20% of individuals with positive PCR would not have been detected if the criteria had not been extended to the presence of nonspecific symptoms(13). It also constitutes a control measure that is likely to reduce the anxiety of health personnel, just as measures to reinforce safety and protection in the care of this type of patient do(14). In this sense, local worker protection policies may have influenced both the percentage of sick workers and that of asymptomatic carriers to be small.

Regarding serology, some authors point it out as an important tool in pandemic control(15). Its use has allowed establishing epidemiological relationships between different clusters, since it would identify past infections(16). In any case, adding individuals with positive IgG or IgM as past infections, would represent a small percentage of the total of our sample. However, the sensitivity and specificity of the serology by immunochromatography technique do not seem superior to the immunoassay(17), especially in asymptomatic individuals and in areas of low prevalence, so these results should be interpreted with caution.

A clear limitation of our work is the sensitivity of naso and oropharyngeal PCR, limited to less than 70% in admitted symptomatic patients(18). There is the theoretical possibility that there was some other undetected carrier, although in any case in our casuistry it should be accidental. It is worth noting that in the areas of greatest exposure (Emergencies, Intensive Care and the CoVID plant) the recruitment of health workers was 100%. Our experience reflects the status of these workers at a certain epidemiological moment, one month after the Spanish Government decreed confinement measures throughout the country, so it is not possible to extrapolate it to other times such as the start of the pandemic in Spain. On the other hand, the study began to be designed in early March, so the presence of anosmia or agueusia was not included in the health questionnaire, but was not included later.

In conclusion, the percentage of asymptomatic healthcare workers with SARSCoV-2 in our hospital center in the units with the highest occupational exposure has turned out to be very low. Hopefully, this information will create reassurance for both healthcare workers and patients.

From the Internal Medicine Unit (JO, MDM, JG), Microbiology Unit (AMC), Laboratory Area (MLH), Preventive Medicine (VF), Occupational Health Department (MJS); Hospital Costa del Sol (Marbella, SPAIN).

## Data Availability

Data are available for investigators after publication

## References

1. Spiteri G, Fielding J, Diercke M, Campese C, Enouf V, Gaymard A, et al. First cases of coronavirus disease 2019 (COVID-19) in the WHO European Region, 24 January to 21 February 2020. Euro Surveill. 2020 Mar;25(9).

2. Wu Z, McGoogan JM. Characteristics of and Important Lessons From the Coronavirus Disease 2019 (COVID-19) Outbreak in China: Summary of a Report of 72314 Cases From the Chinese Center for Disease Control and Prevention. JAMA. 2020 Feb 24.

3. (ISCIII) EC-RCC. Informe nº 25. Situación de COVID-19 en España a 23 de abril de 2020. 2020 [cited 2020 25th april 2020]; Available from: https://www.isciii.es/QueHacemos/Servicios/VigilanciaSaludPublicaRENAVE/EnfermedadesTransmisibles/Documents/INFORMES/Informes%20COVID-19/Informe%20n%C2%BA%2025.%20Situaci%C3%B3n%20de%20COVID-19%20en%20Espa%C3%B1a%20a%2023%20de%20abril%20de%202020.pdf

4. Munster VJ, Koopmans M, van Doremalen N, van Riel D, de Wit E. A Novel Coronavirus Emerging in China – Key Questions for Impact Assessment. N Engl J Med. 2020 Feb 20;382(8):692–4.

5. Li R, Pei S, Chen B, Song Y, Zhang T, Yang W, et al. Substantial undocumented infection facilitates the rapid dissemination of novel coronavirus (SARS-CoV2). Science. 2020 Mar 16.

6. Folgueira MD M-RC, Alonso-López MA, Delgado R on behalf of the Hospital 12 de Octubre COVID-19 Study Groups. SARS-CoV-2 infection in Health Care Workers in a large public hospital in Madrid, Spain, during March 2020. medRxiv; 2020.

7. III IdSC. Situación de COVID-19 en España [Actualizado a 22 de abril de 2020 a las 21:00 horas]. April, 23th 2020 ed. https://covid19.isciii.es 2020.

8. Arons MM, Hatfield KM, Reddy SC, Kimball A, James A, Jacobs JR, et al. Presymptomatic SARS-CoV-2 Infections and Transmission in a Skilled Nursing Facility. N Engl J Med. 2020 Apr 24.

9. Wang D, Hu B, Hu C, Zhu F, Liu X, Zhang J, et al. Clinical Characteristics of 138 Hospitalized Patients With 2019 Novel Coronavirus-Infected Pneumonia in Wuhan, China. JAMA. 2020 Feb 7.

10. Gandhi M, Yokoe DS, Havlir DV. Asymptomatic Transmission, the Achilles’ Heel of Current Strategies to Control Covid-19. N Engl J Med. 2020 Apr 24.

11. Rocklov J, Sjodin H, Wilder-Smith A. COVID-19 outbreak on the Diamond Princess cruise ship: estimating the epidemic potential and effectiveness of public health countermeasures. J Travel Med. 2020 Feb 28.

12. Tong ZD, Tang A, Li KF, Li P, Wang HL, Yi JP, et al. Potential Presymptomatic Transmission of SARS-CoV-2, Zhejiang Province, China, 2020. Emerg Infect Dis. 2020 May;26(5):1052–4.

13. Chow EJ, Schwartz NG, Tobolowsky FA, Zacks RLT, Huntington-Frazier M, Reddy SC, et al. Symptom Screening at Illness Onset of Health Care Personnel With SARS-CoV-2 Infection in King County, Washington. JAMA. 2020 Apr 17.

14. Schwartz J, King CC, Yen MY. Protecting Health Care Workers during the COVID-19 Coronavirus Outbreak –Lessons from Taiwan’s SARS response. Clin Infect Dis. 2020 Mar 12.

15. Winter AK, Hegde ST. The important role of serology for COVID-19 control. Lancet Infect Dis. 2020 Apr 21.

16. Yong SEF, Anderson DE, Wei WE, Pang J, Chia WN, Tan CW, et al. Connecting clusters of COVID-19: an epidemiological and serological investigation. Lancet Infect Dis. 2020 Apr 21.

17. Clínica SEdEIyM. Recomendaciones de SEIMC sobre el uso de las pruebas de detección de anticuerpos. April, 27th 2020 ed. https://seimc.org/contenidos/documentoscientificos/recomendaciones/seimc-rc-2020-Recomendaciones_uso_de_las_pruebas_de_deteccion_de_anticuerpos.pdf 2020.

18. Wang W, Xu Y, Gao R, Lu R, Han K, Wu G, et al. Detection of SARS-CoV-2 in Different Types of Clinical Specimens. JAMA. 2020 Mar 11.

